# Differential Network-Based Causal Graph Learning for Cardiovascular Recurrence Risk Prediction and Factor Discovery

**DOI:** 10.64898/2026.03.16.26348547

**Authors:** Mengyao Zhou, Ming Zhang, Jingjia Wang, Chunli Shao, Guiying Yan

**Author notes:** (Corresponding author: Guiying Yan and Chunli Shao.). (Mengyao Zhou and Ming Zhang contributed equally to this work.). This work has been submitted to the IEEE for possible publication. Copyright may be transferred without notice, after which this version may no longer be accessible.

## Abstract

Cardiovascular disease is one of the leading causes of death worldwide, with myocardial infarction (MI) being a major cause of both morbidity and mortality among cardiovascular patients. MI Patients face a higher risk of cardiovascular disease recurrence afterwards. Therefore, accurately predicting the risk of recurrence and identifying key risk factors are crucial for clinical decision-making. In this paper, we consider the interrelationships among cardiovascular factors from a systemic perspective. We first construct a differential network for each patient to capture individual-specific deviations in factor relationships and propose a novel method, termed Causal Factor-aware Graph Neural Network (CFGNN), which integrates factor interactions to predict the recurrence risk of MI patients while uncovering key risk factors from a causal perspective. Experimental results demonstrate that CFGNN performs well on hospital-derived datasets in real world, effectively identifying several key risk factors. This method not only deepens our understanding of cardiovascular disease, but also paves the way for more targeted and effective interventions.

## I. Introduction

Cardiovascular disease is a major global public health problem and one of the leading causes of mortality, imposing a substantial health and economic burden on populations world-wide. In recent years, the incidence of cardiovascular disease has been steadily rising and is predicted to continue increasing in the future [1]. Among these, myocardial infarction (MI) is particularly life-threatening, resulting in myocardial cell necrosis [2]–[4]. Although advancements in MI treatment have significantly reduced mortality rates [5]–[7], these patients face a high risk of recurrent cardiovascular disease (Fig. 1), such as heart failure, recurrent myocardial infarction, and arrhythmias [8]. These complications not only substantially increase patient mortality but also severely impact quality of life and impose additional strain on healthcare systems. Therefore, accurately predicting long-term recurrence risk, identifying key risk factors, and guiding effective clinical interventions are essential for improving patient outcomes and optimizing healthcare resource allocation [9].

**Fig. 1.**
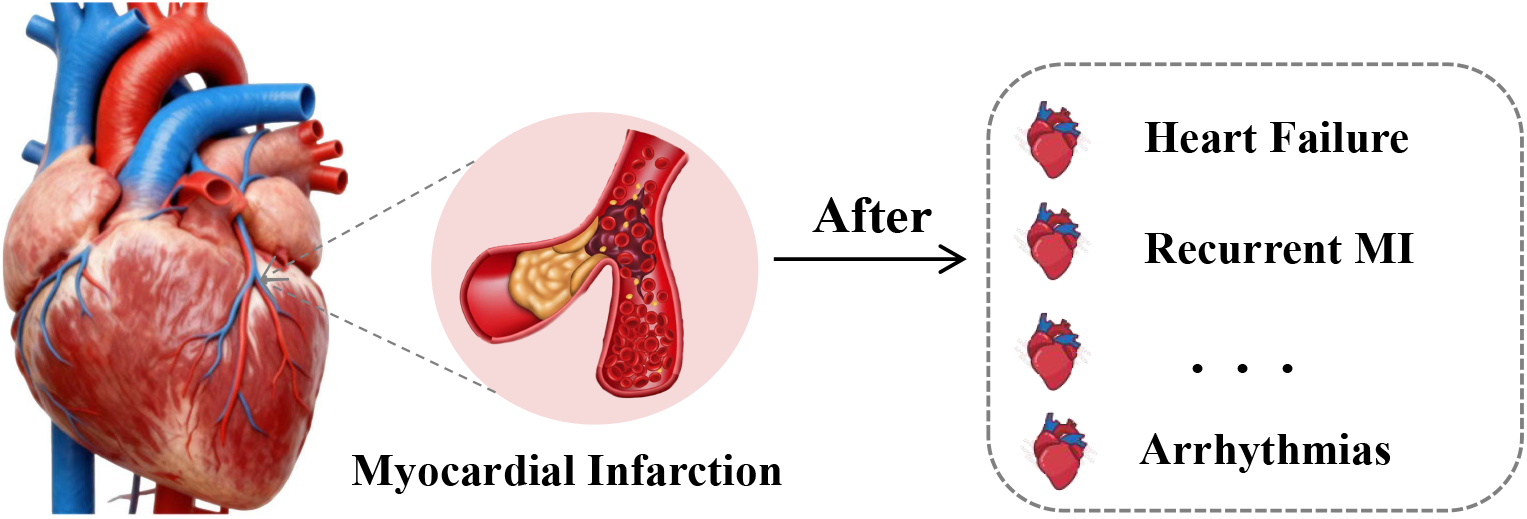
Myocardial infarction Patients face a high risk of recurrent cardiovascular diseases.

Existing methods for predicting the recurrence risk of cardiovascular disease mainly rely on traditional statistical techniques or machine learning models. However, traditional statistical approaches often struggle to efficiently process large-scale datasets and may not scale well with the growing complexity of clinical data. On the other hand, conventional machine learning methods heavily depend on manual feature engineering, which limits their generalizability and robustness. Furthermore, both approaches typically overlook the intricate interactions among risk factors, which are critical for accurate risk prediction and personalized treatment strategies. In recent years, the rise of network medicine offers a systematic analytical framework that allows researchers to comprehensively explore disease complexity, identify key pathogenic factors [10], and uncover relationships among factors [11]. In the context of cardiovascular disease recurrence prediction, network medicine facilitates a more comprehensive and realistic model by revealing key risk factors and interactions among various factors.

In this paper, we collect and process treatment and follow-up data from real-world myocardial infarction (MI) patients to predict the risk of cardiovascular events recurrence after MI, and also identify key risk factors. Specifically, we first construct a differential network using the clinical data of each MI patient to characterize individual-level factor relationships. To alleviate the class imbalance problem in the data, we introduce the GraphSMOTE method for data augmentation to improve the generalization ability of the model. Subsequently, we propose Causal Factor-aware Graph Neural Network(CFGNN) selects the key recurrence risk factors from a causal perspective for risk prediction. This method not only improves prediction performance compared to traditional methods, but also aids clinical decision-making. The main contributions of this study include the following aspects:

- We construct a high-quality real-world dataset of patients with myocardial infarction through comprehensive data collection, integration, and processing, ensuring the authenticity and reliability of the data.
- We employ network methods to characterize factors and their interactions and introduce GraphSMOTE for data augmentation to address the issue of class imbalance in network data.
- We propose CFGNN that utilizes causal invariance to select key factors influencing recurrence risk, enabling interpretable risk prediction and enhancing the clinical applicability of the model.
- Finally, we conduct extensive experiments on real world datasets, and the results show that CFGNN performs excellently in predicting cardiovascular event recurrence risk, not only improving the precision of prediction compared to traditional methods, but also successfully identifying some key risk factors.

## II. Related Work

Existing methods for predicting recurrent cardiovascular events and identifying key risk factors are mainly based on classical statistical approaches, typically involving the establishment of statistical models or scoring systems for risk analysis. Wilson et al. [12] developed a prediction model based on traditional risk factors and found that patients with multiple vascular diseases had a higher risk of cardiovascular event recurrence. Mureddu et al. [13] used a statistical model to show that residual high thrombotic risk significantly increases the likelihood of recurrent hospitalization after myocardial infarction. Yeo et al. [14] applied statistical methods and identified age and diabetes as major predictors of recurrent vascular events and mortality across different populations. Wu et al. [15] used a Cox regression model to identify risk factors for new-onset atrial fibrillation after myocardial infarction, highlighting age, low ejection fraction, and poor nutrition as key contributors. However, these statistical methods are heavily based on initial assumptions, making it difficult to fully capture the complex interactions in high-dimensional biomedical data.

In recent years, there has been increasing attention on using mathematical models and machine learning methods to predict the risk of disease recurrence after myocardial infarction. Aziz et al. [16] employed feature selection methods combined with artificial intelligence algorithms to predict patient mortality, so as to optimize the classification effect of patients with ST-segment elevation myocardial infarction. Yang et al. [17] combined machine learning with SHAP values to improve both prediction and interpretability. SHAP analysis helped identify important risk factors such as age, blood pressure, and lipid levels, making the model more useful for clinical decision-making. Khanet al. [18] employed a variety of machine learning methods were used to predict the risk of recurrent cardiovascular diseases in the Pakistani population, and it was found that random forest was the best model adapted to this data. These studies demonstrate that machine learning methods hold significant potential for cardiovascular disease risk prediction. However, they still face limitations, including a strong reliance on manual feature extraction and a limited ability to capture complex interactions among risk factors, highlighting the need for further improvements and optimization.

Over the past few years, network-based approaches have demonstrated significant potential in the medical field [19]. For example, Lin et al. [20] utilized brain network features to improve the prediction of Alzheimer’s disease. Yin et al. [21] proposed a network-based multi-omics integration framework to gain deeper insights into disease pathogenesis. While network methods are highly effective at integrating complex biological relationships (e.g., genes, diseases) and improving prediction [21], their application in predicting cardiovascular event recurrence remains under-explored. Therefore, this work focuses on integrating multi-source clinical data and quantifying interactions among risk factors to improve the accuracy of recurrent cardiovascular event prediction.

## III. Basic Formulation

### A. Notations

Let *G* = (*V, E, X*) denote a graph, where *V* is the vertex set containing *N* unique vertices and *E* is the edge set containing *M* edges. *X* ∈ *R*^*N*×*d*^ denotes node features matrix with dimension *d* and we use one-hot vectors as the features in this paper. *A* ∈ *R*^*N*×*N*^ is the adjacency matrix, the element *a*_*ij*_ = 1 if edge (*v*_*i*_, *v*_*j*_) ∈ *E* exists, and *a*_*ij*_ = 0 otherwise. The degree matrix of *G* is denoted as *D, d*_*ii*_ = ∑_*j*_ *A*_*ij*_ and 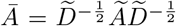 is the regularized adjacency matrix, where *Ã* = *A* + *I* is an adjacency matrix with self-connections and 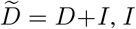 is the identity matrix. 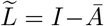 is the Laplacian matrix that represents a normalized symmetric positive semidefinite network.

### B. Graph Neural Networks

Graph Neural Networks (GNNs) leverage both the graph structure and node features *X* to learn a node’s representation vector *h*_*v*_, or the entire graph’s representation *h*_*G*_. Typically, GNNs employ a neighborhood aggregation strategy, where a node’s representation is iteratively updated by aggregating the representations of its neighbors. After *k* iterations, the node’s representation captures the structural information within its *k*-*hop* network neighborhood. Formally, the *k*-*th* layer of a GNN is defined as:

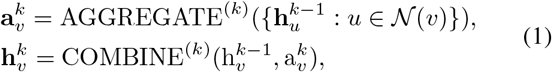

where 𝒩 (*v*) represents the neighbors of node *v*, 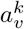 is the representation vector of the neighbor aggregation and 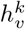 is the representation vector of the node *v* ∈ *V* at *k*-th layer. The choices of AGGREGATE^(*k*)^(*·*) and COMBINE^(k)^(*·*) can be diverse among different GNNs. GCN is one of the most popular GNN structures, which could be viewed as a special case of Eq.(1). Each layer of GCN can be written as:

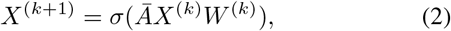

where *X*^(*k*)^ is the representation matrix of the output of the *k*-th layer, *σ* is an activation function.

### C. Problem Definition

Let *P* = {*p*_1_, *p*_2_, *p*_3_…*p*_*m*_}denote the set of MI patients and *F* = {*f*_1_, *f*_2_, *f*_3_…*f*_*n*_} denote the set of factors. Each patient *p*_*i*_ ∈ *P* is associated with a feature vector *X*_*i*_ ∈ *R*^*n*^, each component of *X*_*i*_ corresponds to the value of a factor in *F* .

Our research focuses on the following two problems:

1. **Patient label prediction:** Let *P* ^*lab*^⊆ *P* denote the subset of patients with known labels *y*_*i*_∈ *Y*, and *P* ^*un*^ = *P \P* ^*lab*^ denote the unlabeled subset. The objective is to learn a classification function *f* : *P→ Y*, trained on *P* ^*lab*^, that accurately predicts labels for patients in *P* ^*un*^.
2. **Identify key risk factors:** The objective is to identify a subset of factors *V* ^*\**^ ⊆ *F* that minimizes the prediction error:

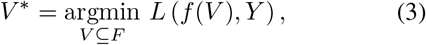

*L*(*·*) is a loss function.

## IV. Model Framework

Based on the perspective of network medicine, we first construct individual-level differential networks to capture interactions among clinical factors. Next, GraphSMOTE is applied to enhance minority-class representation and address class imbalance. Then, we propose a novel model Causal Factor-aware Graph Neural Network (CFGNN) to predict cardiovascular event recurrence in MI patients. The model framework is shown in the Fig. 2.

**Fig. 2.**
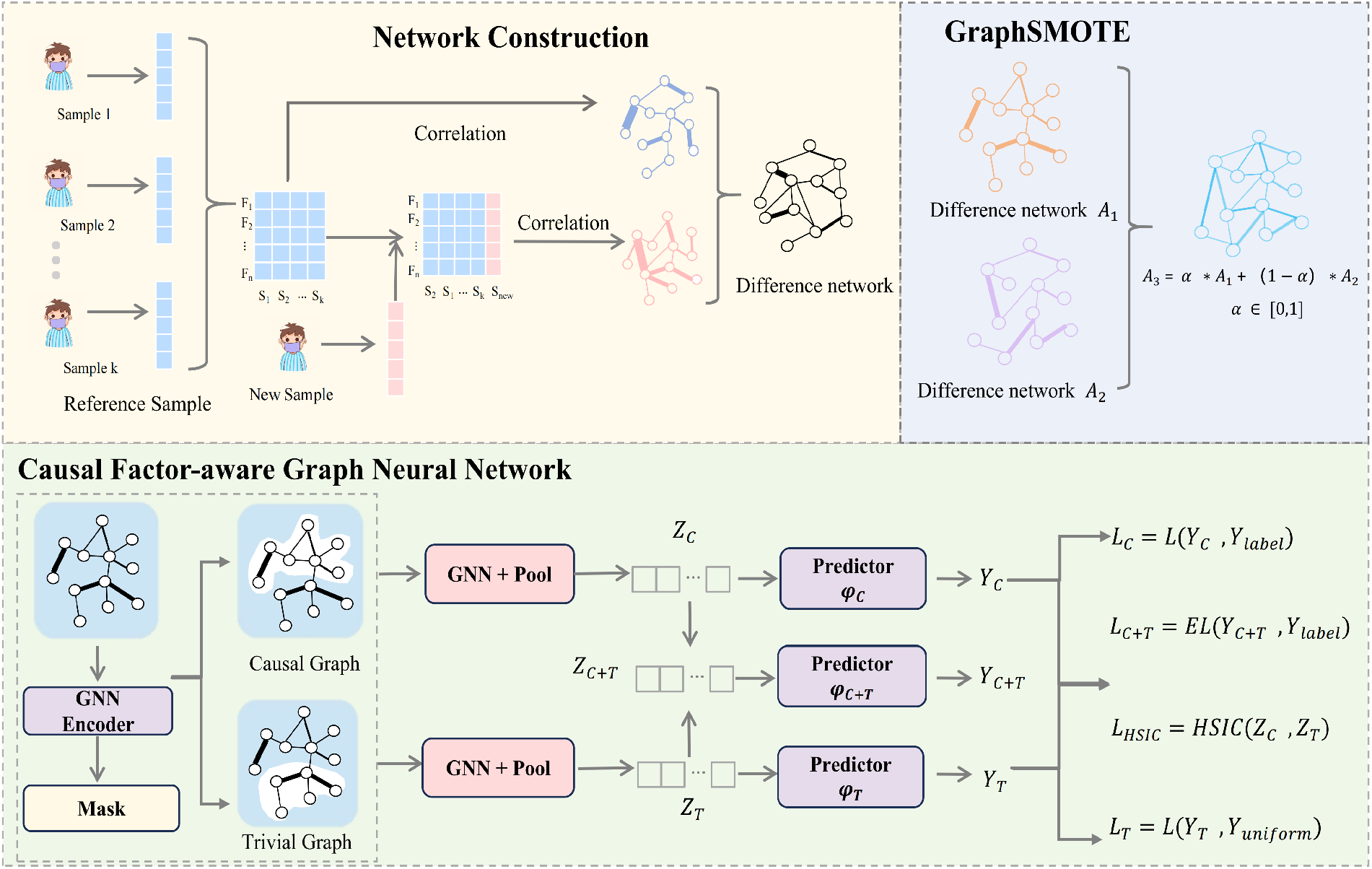
The Flowchart of the method. This figure outlines the proposed pipeline, which consists of three main stages: Network Construction, GraphSMOTE, and the CFGNN Model.

### A. Construct Differential Network from Individual Level

Let *P*_*ref*_ ⊆ *P* be the set of reference samples in MI patients who have not experienced recurrent cardiovascular events afterwards and *F* = {*f*_1_, *f*_2_, *f*_3_…*f*_*n*_} be the set of factors. The Pearson correlation coefficient (PCC) between factors *f*_*i*_ and *f*_*j*_ related to the set *P*_*ref*_ is calculated as:

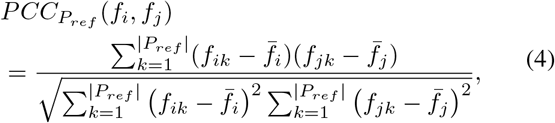

Here, *f*_*ik*_ and *f*_*jk*_ are the expression values for factors *f*_*i*_ and *f*_*j*_ of the *kth* sample in 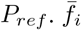 and 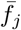 are the average factor-expression values of factors *f*_*i*_ and *f*_*j*_, respectively. After calculating the correlation between every two factors we can get the matrix 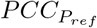.

For each new sample *p*_*l*_ ∈ *P\P*_*ref*_, we add it with the reference samples and a new *PCC* is calculated for every two factors using Eq.(4), we denote this matrix as 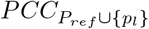 .Subsequently, the difference between these two correlation matrices is computed, which is formally expressed as follows:

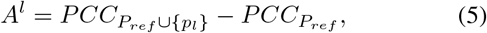

*A*^*l*^ characterizes the difference in factor relationships when sample *p*_*l*_ is added. We compute this matrix for every sample *p*_*l*_ and obtain the set 𝒜= {*A*^*l*^ | *p*_*l*_∈*P\ P*_*ref*_}. Each *A*^*l*^∈ 𝒜corresponds to a difference network *G*^*l*^ where nodes represent factors and edge weights are defined by the correlation differences in *A*^*l*^. We denote the corresponding set of graphs as 𝒢= *G*^*l*^| *p*_*l*_ ∈ *P\P*_*ref*_ . Finally, to reduce the complexity of the data, we refer to [22], [23] and use a testing method to remove edges that do not exhibit significant difference.

### B. GraphSMOTE

In medical data, class imbalance is a prevalent issue that can significantly affect the generalization of the model, particularly leading to bias in predicting results. Previous studies on sequential data imbalance have utilized methods like SMOTE [24]. Here, we extend this principle to the graph domain by proposing GraphSMOTE to mitigate class imbalance in graph-structured data.

Let 𝒢^*lab*^ represent the set of labeled graphs corresponding to labeled patients and 𝒜^*lab*^ represent the set of their associated adjacency matrices. We denote the distance between *A*^*p*^, *A*^*q*^ ∈ 𝒜^*lab*^ as follow:

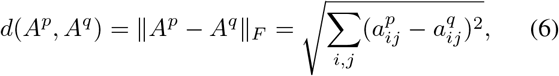

Where 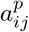 represents the element of the *A*^*p*^. The method first randomly selects a sample *G*^*p*^∈ 𝒢^*lab*^ of minority class and identifies the *K* closest samples of the same class based on the above distance metric. Subsequently, a sample *A*^*q*^ is randomly selected from these K samples and a random number between 0 and 1 is generated to perform linear interpolation, producing a new synthetic sample. The specific formulation is as follows:

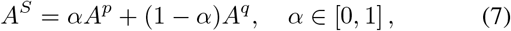

Where *A*^*S*^ represent the newly synthesized sample and we denote the set of labeled graphs after GraphSMOTE as 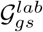 . Through the above steps, GraphSMOTE enhances the representation of the minority class while preserving the structural properties of the graph.

### C. Causal Factor-aware Graph Neural Network

From a causal-based perspective, patient features are categorized as causal features and trivial features. Causal features are those factors that exert a direct, genuine influence on a specific clinical outcome. Therefore, accurately identifying these causal features is important. Based on this insight, we propose Causal Factor-aware Graph Neural Network which screen causal features for improving classification performance. Specifically, we first perform information decomposition, utilizing causal invariance principle to ensure the extracted causal factors accurately reflect the patient’s clinical outcome. We then used this causal information for disease prediction.

#### 1) Decomposition module

Decomposition module aims to decompose the input graph *G* into two subgraphs: the causal subgraph *G*_*c*_ and the trivial subgraph *G*_*t*_. The decomposition module first uses a Graph Neural Network (GNN) to generate a mask matrix *M* ∈ *R*^*n*n*^, where *m*_*ij*_ represents the importance of the edges (*v*_*i*_, *v*_*j*_) in the graph. The specific expression is as follows:

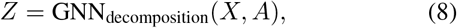

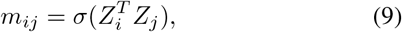

where *X* represents the initial one-hot encoded node features, *σ* is the sigmoid function and *Z* ∈ *R*^*n*d*^ is the representations of the nodes, this process select the edges with the highest masks to construct the *G*_*c*_ and the complement as *G*_*t*_, the specific expression is as follows:

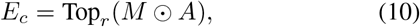

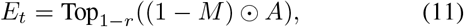

where *E*_*c*_ and *E*_*t*_ represent the edge set of the *G*_*c*_ and *G*_*t*_, respectively; Top_*r*_ selects the *top*-*K* edges with *K* = *r\** |*E*|, r is a hyperparameter, ⊙ is the element-wise product. Having obtained the edge sets, we can distill the nodes appearing in the edges to construct *G*_*c*_ and *G*_*t*_.

#### 2) Prediction module

After decomposition, we use the causal subgraph *G*_*c*_ for disease risk prediction. We also leverage the trivial subgraph *G*_*t*_ during training to ensure that trivial features do not influence the prediction. The specific expression is as follows:

- First, we use GNN to obtain the representations of *G*_*c*_ and then make predictions:

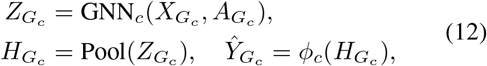

- Then, we use GNN to obtain the representations of *G*_*t*_ and then make predictions:

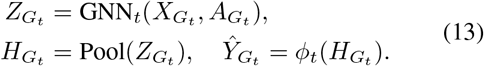

Where 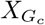 and 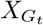 represent the node representation of *G*_*c*_ and *G*_*t*_, 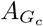and 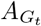 represent the adjacency matrix of *G*_*c*_ and *G*_*t*_, 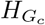 and 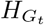 represent the representation of *G*_*c*_ and *G*_*t*_, 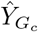 and 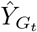 represent the prediction of *G*_*c*_ and *G*_*t*_, *ϕ*_*c*_ and *ϕ*_*t*_ represent classifiers, and here we use multi-layer perceptron as the classifier, Pool is graph pooling operation, here we use mean pooling.

In this section, to ensure the extracted causal subgraph effectively preserves the essential causal information for disease prediction. We use 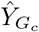 as the result of the classification. Specifically, we designed the causal loss to maximize the contribution of the causal subgraph in the disease classification task, thereby improving both prediction performance. The causal loss function expression for the classification is as follow:

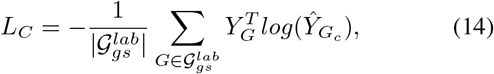

where *Y*_*G*_ represent the true label of the sample *G* and 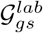 represent the labeled graph set.

To ensure that trivial information, which does not contribute to the prediction, is excluded from the model, a trivial loss is introduced to constrain the trivial subgraph and minimize its contribution to disease prediction. The expression is as follows:

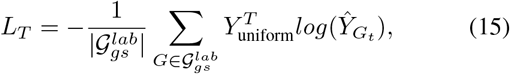

where *Y*_uniform_ represent the uniform label, for data with *c* classes, it is a c-dimensional vector, where each dimension has a value of 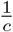 . Trivial loss regularizes the trivial subgraph by minimizing its predictive power, ensuring that the model relies primarily on causal features for decision-making.

Meanwhile, inspired by [25], we design the HSIC loss to measure the independence between the causal and trivial subgraphs. The expression of the HSIC loss is as follows:

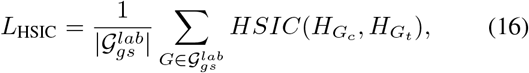

where HSIC is a statistical measure and the specific expression is as follows:

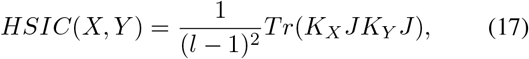

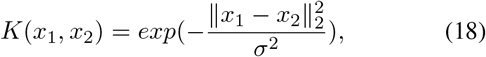

Here, 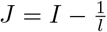, *I* is the *l*-*th* order identity matrix, and *l* is the dimension of the vector *X*.

#### 3) Causal intervention module

In cardiovascular disease risk prediction, patient data often come from diverse clinical backgrounds, leading to distribution differences that pose a significant challenge for model robustness. To mitigate the impact of these distribution discrepancies, and inspired by the principles of causal invariance [26], [27], we introducing a causal intervention module. This principle asserts that a true causal relationship remains stable and consistent across different environments. So, we use classifier to obtain the representations of *G*_*c*_ under different *G*_*t*_. For the *G*_*c*_ of *G*, We combine it with all 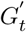 of 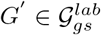, the expression is as follows:

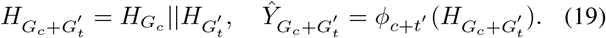

Here, || represents the concatenation operation. Then, causal invariance loss is designed to ensure that the extracted causal subgraph remains stable across different patient populations and clinical environments. The expression is as follows:

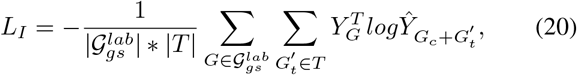

Here, *T* represents the trivial graph set. This ensures reliable risk prediction across different healthcare settings, making it more effective for real-world applications. The total loss function of the model is expressed as follows:

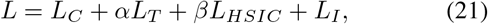

where *α, β* are hyperparameters.

## V. Experiment

### A. Experimental Setting

#### 1) Datasets

To strictly evaluate the generalization capability of our method, we perform experiments on two distinct myocardial infarction (MI) datasets. The first dataset, denoted as MID-I, was collected from Peking University Third Hospital and Fuwai Hospital. It initially comprised clinical data from over 3,000 patients diagnosed with myocardial infarction. The second dataset, denoted as MID-II, is derived from the study in [28].

Both datasets have been anonymized and de-identified to ensure the protection of personally identifiable information (PII). They encompass a diverse range of features, including demographic information (e.g., sex, age, BMI), treatment details (e.g., intraoperative medications, PCI procedures), and follow-up record, due to some limitations, we were only able to acquire partial records for MID-II.

The prediction target for both datasets is consistent. Patient labels are defined based on a two-year follow-up period: patients who experienced a recurrence of cardiovascular disease within two years are labeled as positive samples, while those with no recurrence are labeled as negative samples. To ensure consistency in our analysis, an identical data processing pipeline was applied to both the MID-I and MID-II datasets. The procedure consists of two main stages:

1. Data cleaning and filtering: This step involved removing outliers, correcting inconsistencies, and excluding unreliable or irrelevant records to guarantee the quality of the input data.
2. Feature Normalization: To enhance analytical utility, continuous variables were discretized, and categorical variables were converted into binary indicators. Taking the *Age* variable as an example, it was divided into three distinct factors: age *≤*44, age 45 *™* 59, and age *≥* 60. A patient aged 45 would be assigned a value of 1 for the “age 45*™*59” factor and 0 for the others. This logic was applied uniformly across both datasets to align the feature space. The statistics of the processed datasets are summarized in Table I.

**TABLE I.** Details of the Preprocessed Datasets.

| Dataset | Source | Patients | Features | Classes |
| --- | --- | --- | --- | --- |
| MID-I | PKU & Fuwai | 1,649 | 202 | 2 |
| MID-II | CatLet Study Cohort | 955 | 48 | 2 |

#### 2) Implementation details

We report the mean value and standard deviation of the 10-fold cross-validation results for patient classification. The CFGNN model employs a two-layer Graph Convolutional Network (GCN) as its GNN encoder, utilizing mean graph pooling for feature aggregation. In our experiments, we set the number of reference samples to 90 and the class imbalance neighbor parameter (*K*) to 5. Following the methods in [26], [27], we adaptively adjust the loss function of the causal intervention module, with the loss weighting coefficients *α* and *β* are selected from the set {0.1, 0.2, 0.3, 0.4, 0.5} and {0.001, 0.002, 0.003, 0.004, 0.005} to balance different loss terms. The causal rate *r* is selected from the set {0.5, 0.6, 0.7, 0.8, 0.9} .

#### 3) Baselines

We compare the following representative models with our models.

- Logistic Regression [29]: Logistic Regression is a supervised learning algorithm that classifies data by modeling the probability of class membership using the logistic function.
- KNN [30]: The principle behind KNN is that the class of a sample is determined by the majority class of its k nearest neighbors.
- SVM [31]: SVM is a supervised learning algorithm that classifies data by finding the optimal hyperplane in a high-dimensional space.
- MLP [32]: MLP is a feedforward neural network with multiple layers that learns non-linear input-output mappings.
- KAN [33]: KAN is a neural model using learnable 1D functions instead of fixed activations, improving interpretability and approximation.
- GCN [34]: GCN is a graph neural network that aggregates features from neighboring nodes using spectral-based convolutions.
- GAT [35]: GAT is a graph neural network that uses attention mechanisms to assign different weights to neighbors during feature aggregation.
- GraphSAGE [36]: GraphSAGE is a GNN framework that samples and aggregates information from a node’s local neighborhood to generate embeddings.
- CIN [37]: CIN is a graph model that captures high-order interactions by combining structural and feature-level information through cross-layer integration.
- LEGCN [38]:LEGCN is an extension of GCN that incorporates edge label information to enhance message passing and representation learning.
- GNNExplainer [39]: GNNExplainer is a model that provides interpretability for Graph Neural Networks (GNNs) by identifying the important subgraphs and node features that contribute most to the model’s predictions.
- ERM [40]: This strategy optimizes the model parameters through empirical risk minimization.
- DIR [27]: DIR captures invariant features by minimizing the prediction variance after different interventions.

#### 4) Evaluation metrics

To evaluate the effectiveness of the model, we assess it using the following metrics: Accuracy (ACC), Precision (PRE), Recall (REC), F1-Score, and AUC (Area Under the Curve). These metrics provide a comprehensive measure of the model’s performance from different perspectives.

### B. Comparison result

To validate the effectiveness of the proposed method, a comprehensive comparative analysis was conducted on the MID-I and MID-II, as summarized in Table II and Table III, respectively. Furthermore, the multi-metric performance comparison is visually illustrated via radar charts in Fig. 3. The details of the comparative experiments are as follows:

- First, We utilize SMOTE to balance the dataset and performed classification using various common machine learning models, sequence models and graph-based models.
- In addition, we constructed differential networks, applied GraphSMOTE to address class imbalance, and then performed patient classification using a basic graph-based models.

**TABLE II.**
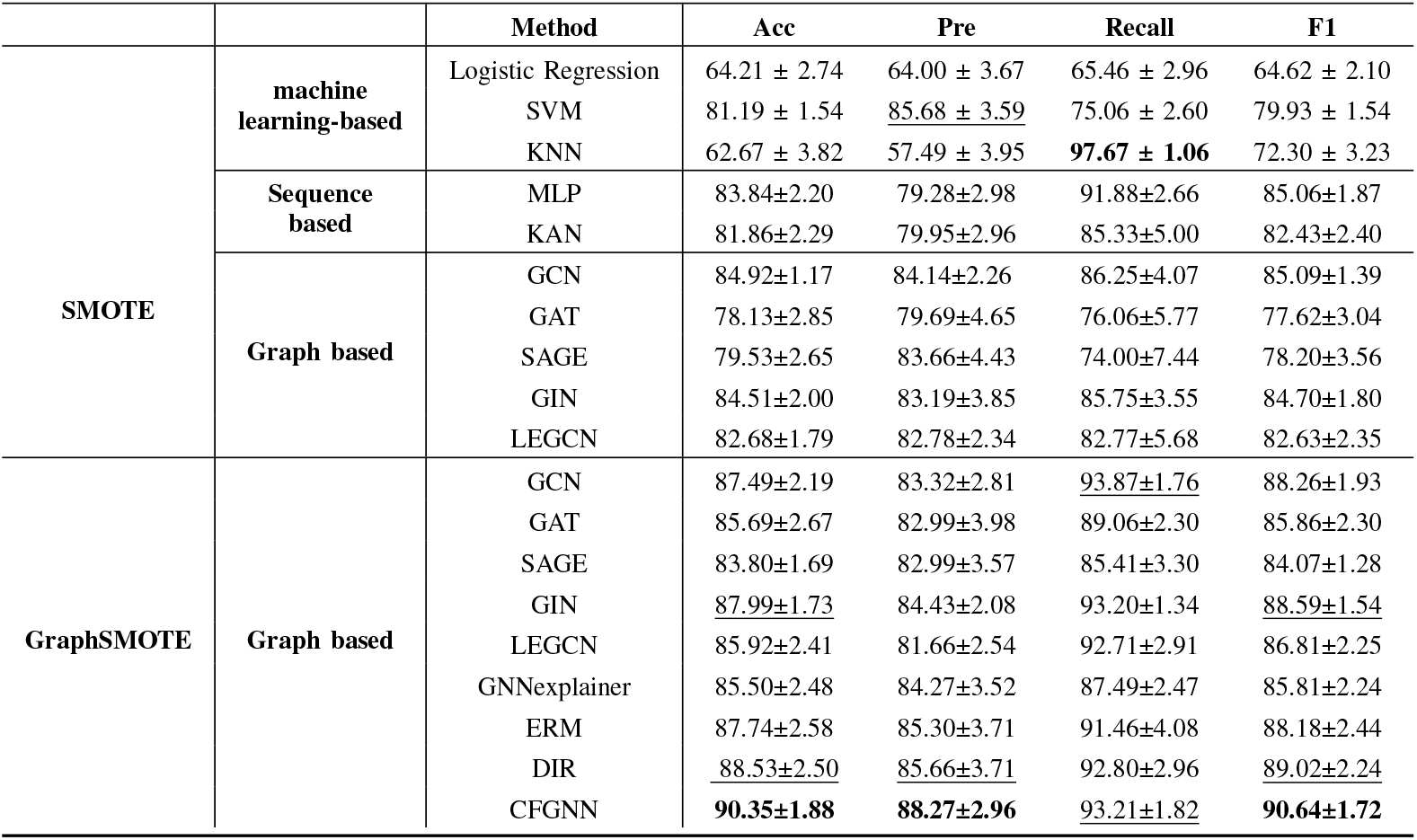
Multi-metric classification results on MID-I. The best results are highlighted in bold, while the second or third best results are indicated with underlined text.

**TABLE III.** Multi-metric classification results on MID-II. The best results are highlighted in bold, while the second or third best results are indicated with underlined text.

|  |  | Method | Acc | Pre | Recall | F1 |
| --- | --- | --- | --- | --- | --- | --- |
| SMOTE | machine learning-based | Logistic Regression | 64.01 ± 3.15 | 45.73 ± 4.33 | 55.97 ± 11.42 | 50.07 ± 7.23 |
|  |  | SVM | 62.26 ± 3.66 | 43.22 ± 5.68 | 47.84 ± 11.32 | 44.98 ± 7.21 |
|  |  | KNN | 57.28 ± 4.99 | 39.70 ± 5.51 | 59.11 ± 10.93 | 47.42 ± 7.30 |
|  | Sequence based | MLP | 68.17 ± 4.38 | 53.21 ± 9.87 | 51.06 ± 16.02 | 49.94 ± 9.77 |
|  |  | KAN | 66.19 ± 3.31 | 52.30 ± 13.09 | 41.12 ± 17.97 | 42.25 ± 10.42 |
|  | Graph based | GCN | 68.28 ± 7.59 | 52.55 ± 8.96 | 67.21 ± 6.10 | 58.40 ± 5.63 |
|  |  | GAT | 68.44 ± 7.74 | 53.03 ± 9.10 | 64.62 ± 8.02 | 57.52 ± 5.43 |
|  |  | SAGE | 65.09 ± 14.97 | 52.88 ± 11.86 | 71.31 ± 16.01 | 58.22 ± 4.74 |
|  |  | GIN | 67.15 ± 8.86 | 51.77 ± 9.88 | 71.13 ± 7.51 | 59.05 ± 6.09 |
|  |  | LEGCN | 69.57 ± 8.28 | 54.79 ± 9.31 | 67.87 ± 10.47 | 59.64 ± 4.00 |
| GraphSMOTE | Graph based | GCN | 70.67 ± 5.12 | 54.96 ± 4.91 | 65.27 ± 7.48 | 59.25 ± 3.30 |
|  |  | GAT | 68.16 ± 6.48 | 52.00 ± 6.23 | 66.07 ± 11.42 | 57.48 ± 4.71 |
|  |  | SAGE | 69.05 ± 6.07 | 53.13 ± 7.16 | 67.64 ± 11.21 | 58.70 ± 5.15 |
|  |  | GIN | 69.95 ± 4.53 | 53.00 ± 5.86 | <u>72.46 ± 9.63</u> | 60.93 ± 6.03 |
|  |  | LEGCN | 70.94 ± 5.94 | 54.87 ± 7.66 | <u>69.09 ± 8.81</u> | 60.79 ± 6.52 |
|  |  | GNNexplainer | 71.40 ± 4.51 | 55.49 ± 5.33 | 66.38 ± 7.68 | 60.13 ± 4.71 |
|  |  | ERM | <u>71.75 ± 5.50</u> | <u>56.30 ± 8.76</u> | 69.04 ± 9.51 | <u>61.36 ± 6.36</u> |
|  |  | DIR | <u>71.70 ± 5.02</u> | <u>56.13 ± 7.88</u> | 63.82 ± 6.72 | <u>59.53 ± 6.52</u> |
|  |  | CFGNN | **72.58 ± 6.68** | **56.76 ± 9.44** | **73.76 ± 6.50** | **63.86 ± 7.42** |

**Fig. 3.**
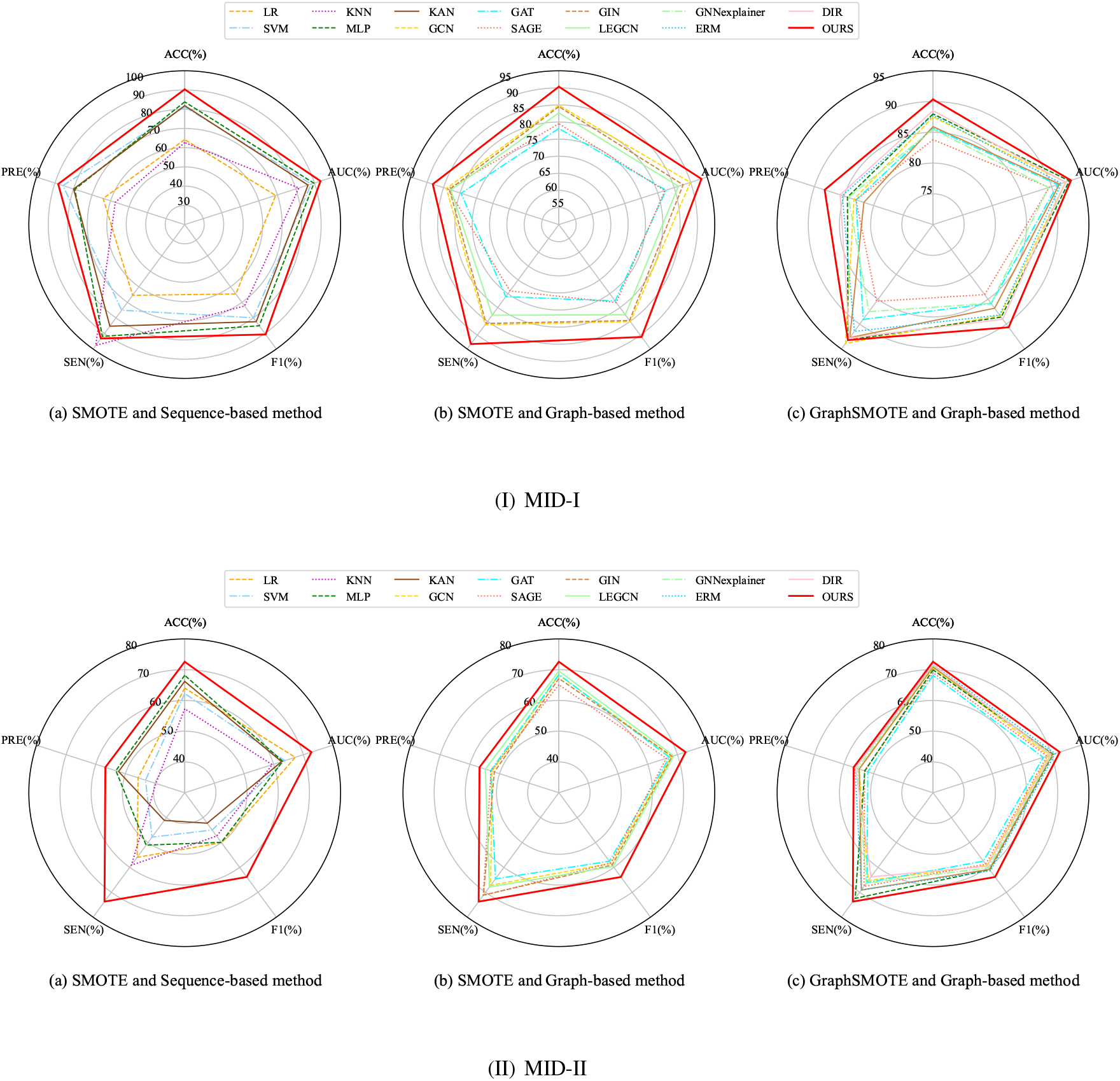
Multi-metric radar chart on MID-I and MID-II: (a) shows the results of sequence-based methods (SMOTE); (b) shows the results of graph-based methods (SMOTE); (c) shows the results of graph-based methods (GraphSMOTE).

The results demonstrate that, following the application of SMOTE, graph-based classification models outperform sequence-based models. This highlights the effectiveness of the differential network in capturing informative structural representations. Moreover, we compared the performance of graph classification under two resampling strategies, GraphSMOTE and traditional SMOTE, thereby providing additional validation of the superiority of GraphSMOTE in handling class imbalance in graph-structured data. Finally, a comparison between the proposed CFGNN and other interpretable graph classification models reveals that CFGNN achieves superior performance across multiple evaluation metrics.

It is also worth noting that the overall predictive performance on MID-I consistently surpasses that on MID-II. This disparity is primarily attributed to the fact that MID-I encompasses more comprehensive patient information, which provides a richer basis for classification.

### C. Ablation study

We perform ablation studies on the MID-I and MID-II to evaluate the individual contributions of different constraints in our models. The specific observations are as follows:

- **w/o** *L*_*T*_ : This variant of CFGNN disables the loss *L*_*T*_, performing classification solely on *L*_*C*_, *L*_*HSIC*_ and *L*_*I*_ .
- **w/o** *L*_*HSIC*_: In this CFGNN variant, the restriction *L*_*HSIC*_ is removed, and graph classification is is performed exclusively with *L*_*C*_, *L*_*T*_ and *L*_*I*_ .
- **w/o** *L*_*I*_ : In this CFGNN variant, the *L*_*I*_ constraint is excluded, and graph classification is performed using only the other loss functions.

As shown in Fig. 4, all variants exhibit degraded performance in label prediction, underscoring the importance of these components in enhancing the modeling of interaction types.

**Fig. 4.**
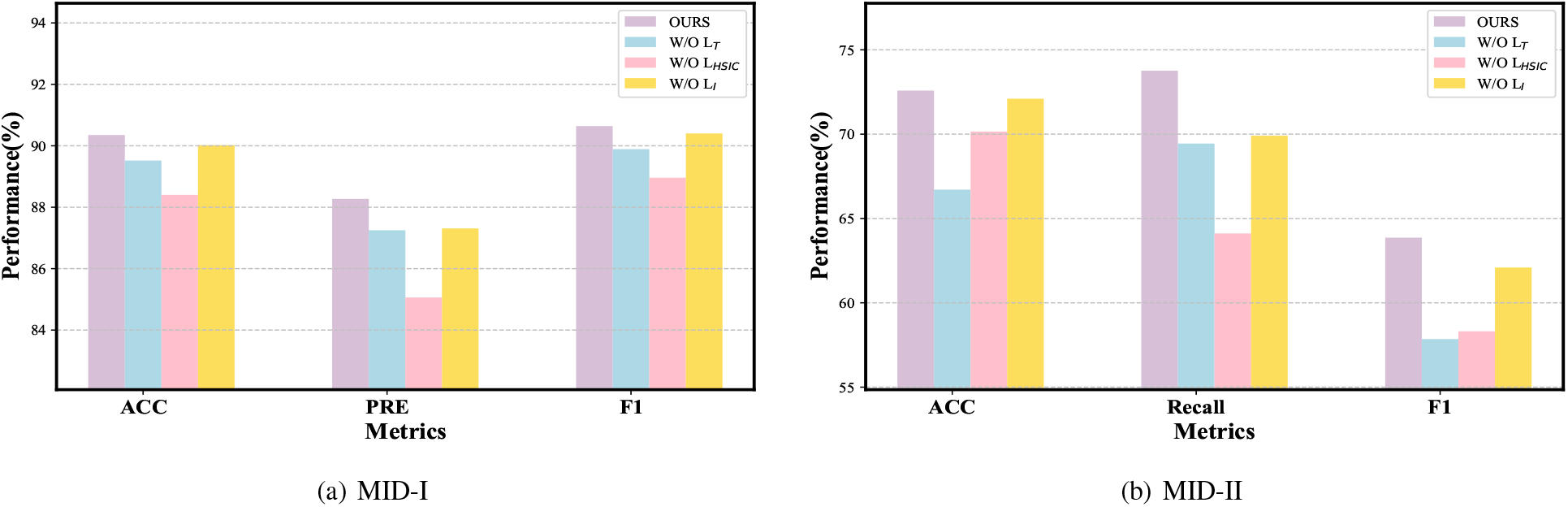
(a) Ablation Study on the MID-I. (b) Ablation Study on the MID-II.

### D. Parameter analysis

In this section, we analyze the hyperparameters of CFGNN on the MID-I, with the specific results shown in the Fig. 5. From the Fig. 5(a), we observe that the classification accuracy first increases and then decreases as the causal ratio *r* changes, peaking at 0.9. This indicates that a causal ratio of 0.9 accurately captures the number of causal factors in the data. When the ratio is too low, some causal factors are excluded, and when it is too high, some non-causal factors may be mixed into the causal subgraph, which aligns with the actual situation. Additionally, Figs. 5(b) and 5(c) show how the classification accuracy changes with variations in the coefficients of the loss function. From these figures, we can see that fluctuations in the coefficients have little impact on the experimental results, reflecting the stability of the model.

**Fig. 5.**
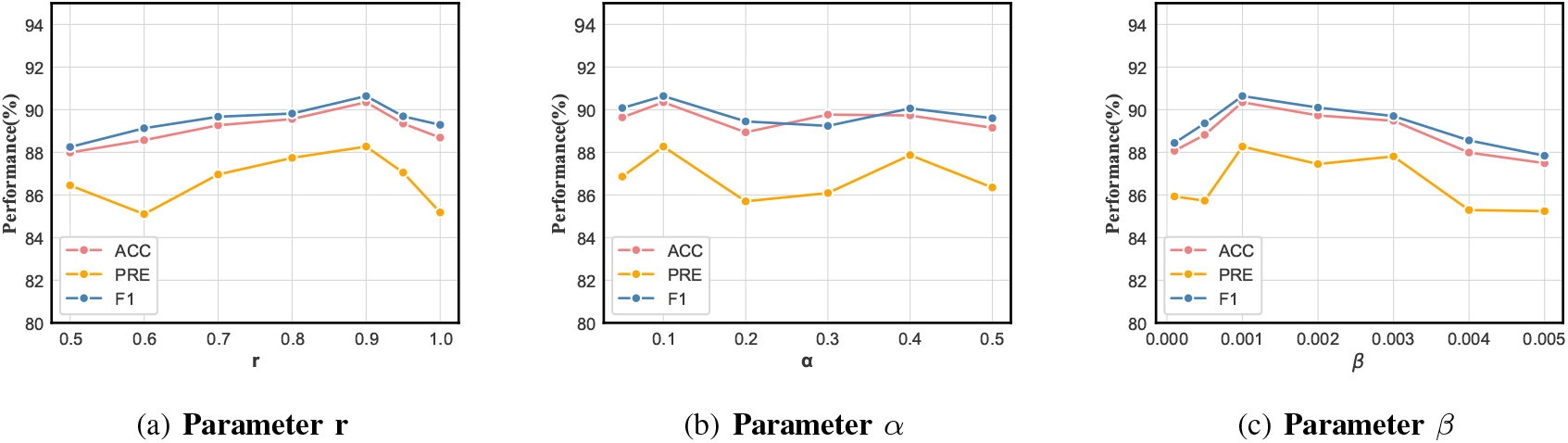
Parameter sensitivity analysis:(a) shows multi-metric results on MID with different parameter r. (b) shows multi-metric results on MID with different parameter ***α***.(c) shows multi-metric results on MID with different parameter ***β***.

### E. Key factor analysis

In this section, we analyze the causal information extracted from on the MID-I by the model. We focus on this dataset specifically due to its comprehensive and rich nature, which provides a more robust basis for interpreting the causal mechanisms. First, we extracted the causal subgraphs for all patients in the test set and counted the frequency of each factor appearing in these causal subgraphs. We then merged the frequencies of different expressions referring to the same factor and ranked the factors accordingly. The specific results are shown in Table IV, we present the top twenty factors from the experimental results of all patients. Through a literature review, we found that sixteen of these twenty factors have been confirmed as risk factors in relevant studies. This further validates the effectiveness of the CFGNN model and enhances the interpretability of key risk factor identification.

**TABLE IV.**
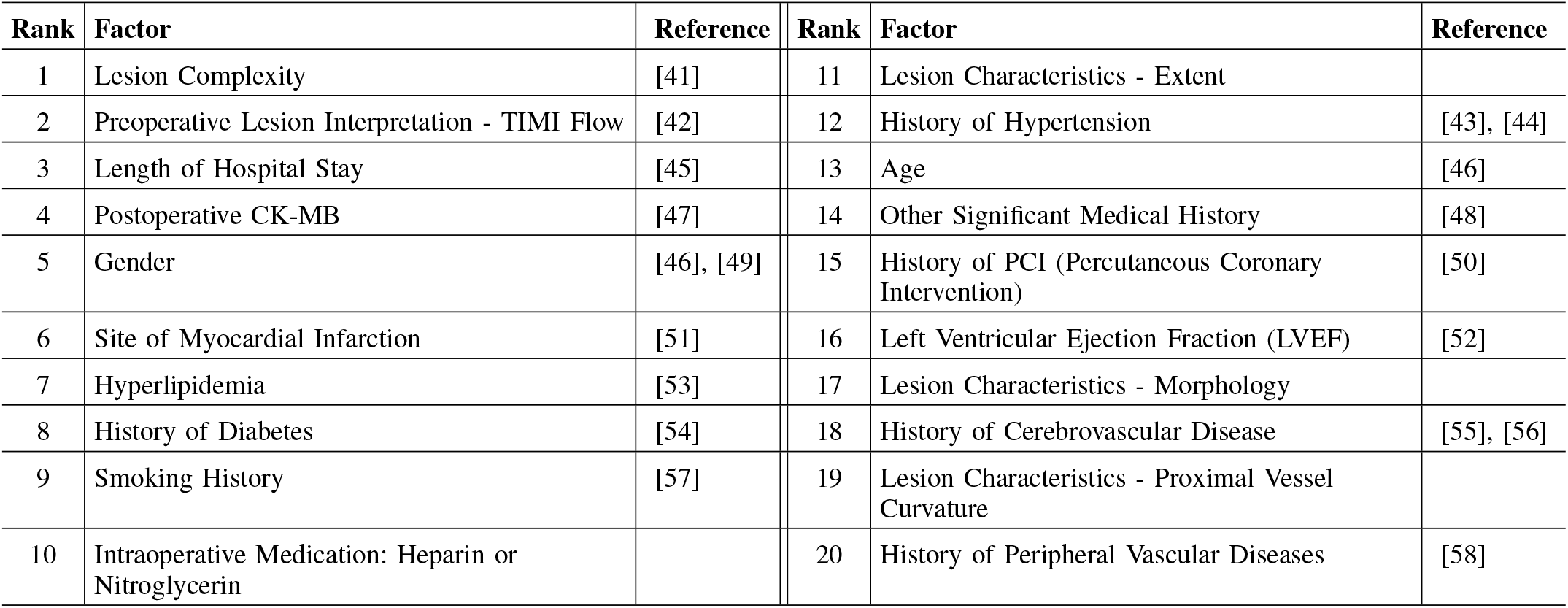
Top 20 key risk factors.

As shown in Table IV, in addition to traditional risk factors such as age and medical history, our study highlights the significant impact of lesion complexity and specific lesion characteristics on recurrent cardiovascular events. Notably, lesion complexity emerged as the most significant factor. This comprehensive metric was synthesized during data collection by integrating the syntax score, degree of bifurcation, severity of occlusion, calcification status, and the number of involved vessels. Our results demonstrate that while traditional factors—such as age and medical history—hold substantial importance in the recurrence of cardiovascular events, specific lesion characteristics, particularly their structural complexity and morphological details, play a more decisive role in identifying high-risk patients. This finding provides clinicians and patients with a new perspective, suggesting that when assessing post-infarction cardiovascular risk, attention should not only be paid to conventional risk factors but also to a comprehensive evaluation of individualized lesion characteristics, thereby enabling the development of more targeted secondary prevention strategies.

### F. Gender-Specific Risk Factor Analysis

In this study, we conducted a gender–specific exploration of the factors affecting the risk of cardiovascular disease recurrence. By comparing the characteristics of female and male patients, differences in the composition of risk factors were identified. Overall, most of the important features selected by gender were also included in the important factors of all patients. The results are shown in Fig. 6.

**Fig. 6.**
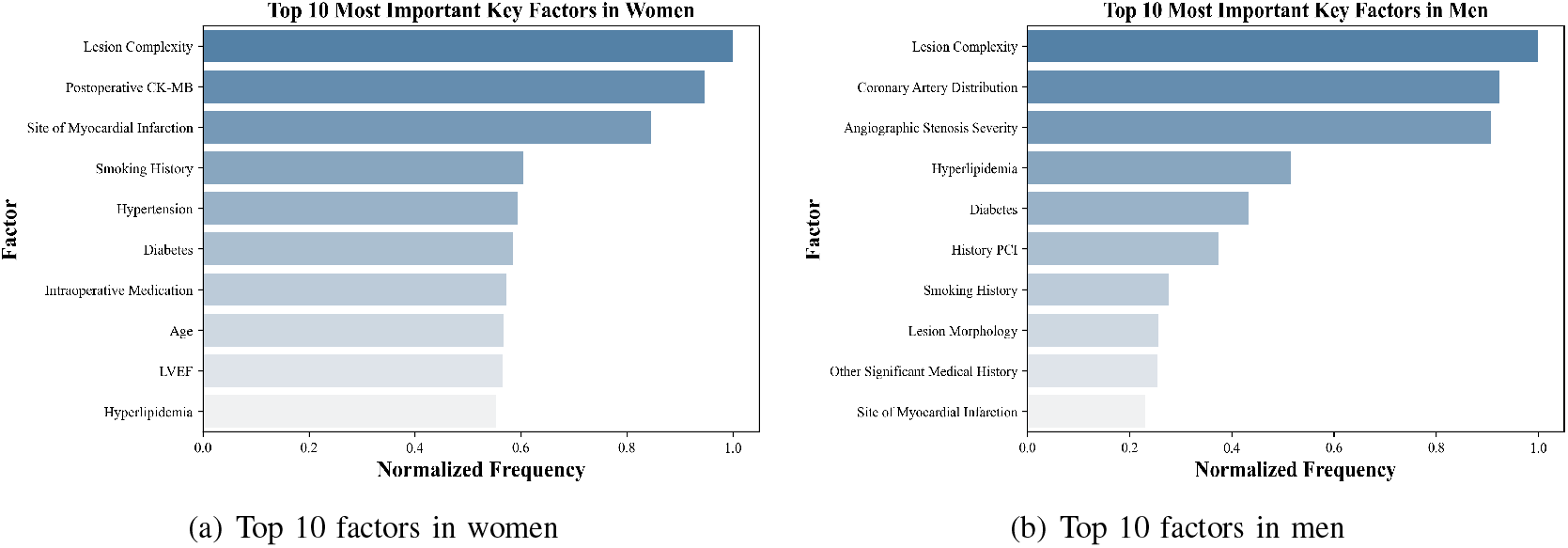
(a) shows the Top 10 factors in women. (b) shows the Top 10 factors in men.

As illustrated in Fig. 6, the risk factors for recurrent cardiovascular disease following myocardial infarction in women tend to be associated with microvascular dysfunction, endothelial impairment, non-obstructive coronary artery disease, postoperative indicators, residual ischemia, and metabolic disorders. This suggests that secondary prevention for female patients should place greater emphasis on postoperative monitoring, metabolic syndrome management, and microvascular protection.

In contrast, the risk factors for men are more concentrated on macrovascular lesions, high plaque burden, and traditional obstructive atherosclerosis, as well as stenosis severity, lesion morphology, lipid levels, and smoking status. Consequently, management strategies for men should prioritize lesion morphology assessment, intensive lipid control, and smoking cessation. These findings are consistent with previously reported literature [59], [60].

### G. Case study

In this section, four representative cases were selected for analysis, and the results are shown in Fig. 7. Figs. 7(a) and 7(b) illustrate female patients without and with recurrent cardiovascular events, respectively, while Figs. 7(c) and 7(d) depict the corresponding scenarios for male patients. Each subfigure represents an individualized differential network constructed for a single patient, reflecting the interaction patterns among clinical and lesion-related factors.The pink nodes highlight the key factors identified through our method, indicating variables that play a dominant role in the patient-specific network structure.

**Fig. 7.**
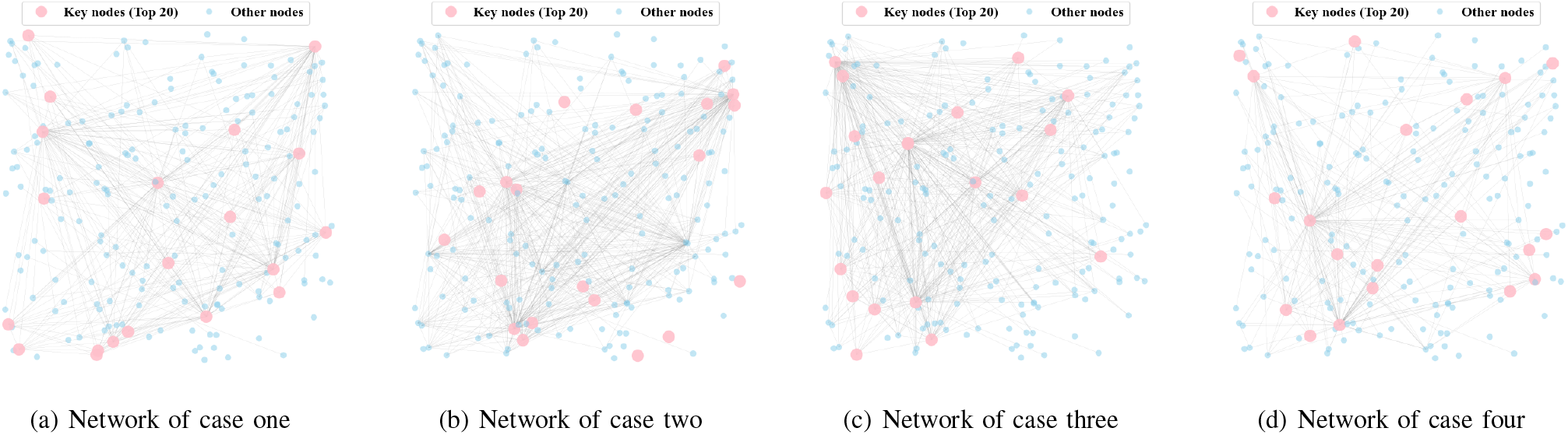
Case-study differential networks.(a) Female patient without recurrent cardiovascular events. (b) Female patient with recurrent cardiovascular events. (c) Male patient without recurrent cardiovascular events. (d) Male patient with recurrent cardiovascular events. Key nodes (highlighted) indicate the most important factors identified by our method.

At the holistic level, although the four cases were constructed based on an identical set of factors, the resulting differential networks exhibited significant individual heterogeneity in terms of connection strength, topological complexity, and the organization patterns of key nodes. These highly inconsistent topological features indicate that factor association patterns at the individual level possess unique complexities that cannot be effectively characterized by a unified group-average structure.

In the analysis of key nodes, the critical factors identified across different cases varied significantly, further underscoring the individualized nature of risk mechanisms. Regarding the female samples, the key nodes for the non-recurrence individual were concentrated on lesion characteristics, postoperative CK-MB, BMI, and the site of infarction. This suggests that the favorable prognosis in this case was primarily associated with the extent of acute myocardial injury and basal metabolic status. Conversely, the key risk factors for the recurrence female sample mainly included LVEF, hypertension, diabetes, history of peripheral vascular disease, and TIMI flow grade. This reflects that the risk of recurrence may be predominantly driven by the burden of chronic underlying diseases and cardiac functional reserve.

A similar pattern of heterogeneous differentiation was observed in the male samples. The key nodes for the non-recurrence male individual primarily involved postoperative hemoglobin, history of CABG, site of infarction, admission blood pressure, and hyperlipidemia. This reflects that the prognostic outcome was more heavily influenced by the combined effects of prior treatment history and perioperative physiological indicators. In contrast, the key factors for the recurrence male sample focused on coronary dominance, lesion characteristics, history of PCI, BMI, and site of infarction, suggesting a close intrinsic correlation between disease recurrence and both coronary anatomical variations and the history of interventional therapy.

This analysis provides personalized risk feature identification and evaluation for myocardial infarction patients, which is useful for achieving refined prognostic prediction and intervention strategy formulation.

## VI Conclusion

In this study, we focused on predicting recurrent cardiovascular events and identifying key risk factors in MI patients. We collected real-world clinical data to construct a high-quality dataset encompassing multi-dimensional patient features. From the perspective of network medicine, we built personalized differential networks for each patient and proposed the GraphSMOTE method to address data imbalance. Building on this foundation, we proposed a Causal Factor-aware Graph Neural Network to identify causal components in MI patient data and achieve more accurate recurrence prediction. Our experimental results show that the proposed model not only outperforms existing approaches in prediction accuracy but also effectively identifies several key recurrence risk factors. In addition to confirming well-known factors, our model uncovers lesion-related features that have drawn attention from clinicians and patients, providing a valuable basis for more precise prognosis and targeted intervention strategies.

## Data Availability

All data produced in the present study are available upon reasonable request to the authors.

## REFERENCES

[1] L.-Y. Ma, W.-W. Chen, R.-L. Gao, L.-S. Liu, M.-L. Zhu, Y.-J. Wang, Z.-S. Wu, H.-J. Li, D.-F. Gu, Y.-J. Yang et al., “China cardiovascular diseases report 2018: an updated summary,” Journal of geriatric cardiology: JGC, vol. 17, no. 1, p. 1, 2020.

[2] M. Saleh and J. A. Ambrose, “Understanding myocardial infarction,” F1000Research, vol. 7, 2018.

[3] G. W. Reed, J. E. Rossi, and C. P. Cannon, “Acute myocardial infarction,” The Lancet, vol. 389, no. 10065, pp. 197–210, 2017.

[4] K. Thygesen, J. S. Alpert, A. S. Jaffe, B. R. Chaitman, J. J. Bax, D. A. Morrow, H. D. White, and E. G. on behalf of the Joint European Society of Cardiology (ESC)/American College of Cardiology (ACC)/American Heart Association (AHA)/World Heart Federation (WHF) Task Force for the Universal Definition of Myocardial Infarction, “Fourth universal definition of myocardial infarction (2018),” Circulation, vol. 138, no. 20, pp. e618–e651, 2018.

[5] E. Puymirat, G. Taldir, N. Aissaoui, G. Lemesle, L. Lorgis, T. Cuisset, P. Bourlard, B. Maillier, G. Ducrocq, J. Ferrieres et al., “Use of invasive strategy in non–st-segment elevation myocardial infarction is a major determinant of improved long-term survival: Fast-mi (french registry of acute coronary syndrome),” JACC: Cardiovascular Interventions, vol. 5, no. 9, pp. 893–902, 2012.

[6] G. Zuanetti, R. Latini, A. P. Maggioni, M. Franzosi, L. Santoro, and G. Tognoni, “Effect of the ace inhibitor lisinopril on mortality in diabetic patients with acute myocardial infarction: data from the gissi-3 study,” Circulation, vol. 96, no. 12, pp. 4239–4245, 1997.

[7] J. A. de Lemos and S. M. Ettinger, “2013 accf/aha guideline for the management of st-elevation myocardial infarction,” Journal of the American College of Cardiology, vol. 61, no. 4, 2013.

[8] K. Smolina, F. L. Wright, M. Rayner, and M. J. Goldacre, “Long-term survival and recurrence after acute myocardial infarction in england, 2004 to 2010,” Circulation: Cardiovascular Quality and Outcomes, vol. 5, no. 4, pp. 532–540, 2012.

[9] G. A. Roth, G. A. Mensah, C. O. Johnson, G. Addolorato, E. Ammirati, L. M. Baddour, N. C. Barengo, A. Z. Beaton, E. J. Benjamin, C. P. Benziger et al., “Global burden of cardiovascular diseases and risk factors, 1990–2019: update from the gbd 2019 study,” Journal of the American college of cardiology, vol. 76, no. 25, pp. 2982–3021, 2020.

[10] K.-I. Goh, M. E. Cusick, D. Valle, B. Childs, M. Vidal, and A.-L. Barabási, “The human disease network,” Proceedings of the National Academy of Sciences, vol. 104, no. 21, pp. 8685–8690, 2007.

[11] F. Barrenas, S. Chavali, P. Holme, R. Mobini, and M. Benson, “Network properties of complex human disease genes identified through genome-wide association studies,” PloS one, vol. 4, no. 11, p. e8090, 2009.

[12] P. W. Wilson, R. D’Agostino Sr, D. L. Bhatt, K. Eagle, M. J. Pencina, S. C. Smith, M. J. Alberts, J. Dallongeville, S. Goto, A. T. Hirsch et al., “An international model to predict recurrent cardiovascular disease,” The American journal of medicine, vol. 125, no. 7, pp. 695–703, 2012.

[13] G. F. Mureddu, C. Greco, S. Rosato, P. D’Errigo, L. De Luca, G. Badoni, P. Faggiano, and F. Seccareccia, “High thrombotic risk increases adverse clinical events up to 5 years after acute myocardial infarction. a nationwide retrospective cohort study,” Monaldi Archives for Chest Disease, vol. 89, no. 3, 2019.

[14] K. K. Yeo, H. Zheng, K. Y. Chow, A. Ahmad, B. P. Chan, H. M. Chang, E. Chong, T. S. J. Chua, D. C. G. Foo, L. P. Low et al., “Comparative analysis of recurrent events after presentation with an index myocardial infarction or ischaemic stroke,” European Heart Journal-Quality of Care and Clinical Outcomes, vol. 3, no. 3, pp. 234–242, 2017.

[15] L. Wu, W. Wang, Y. Gui, Q. Yan, G. Peng, X. Zhang, L. Ye, and L. Wang, “Nutritional status as a risk factor for new-onset atrial fibrillation in acute myocardial infarction,” Clinical Interventions in Aging, pp. 29–40, 2023.

[16] F. Aziz, S. Malek, K. S. Ibrahim, R. E. Raja Shariff, W. A. Wan Ahmad, R. M. Ali, K. T. Liu, G. Selvaraj, and S. Kasim, “Short-and long-term mortality prediction after an acute st-elevation myocardial infarction (stemi) in asians: A machine learning approach,” PloS one, vol. 16, no. 8, p. e0254894, 2021.

[17] F. Yang, Y. Qiao, P. Hajek, and M. Z. Abedin, “Enhancing cardiovascular risk assessment with advanced data balancing and domain knowledge-driven explainability,” Expert Systems with Applications, vol. 255, p. 124886, 2024.

[18] A. Khan, M. Qureshi, M. Daniyal, and K. Tawiah, “A novel study on machine learning algorithm-based cardiovascular disease prediction,” Health & Social Care in the Community, vol. 2023, no. 1, p. 1406060, 2023.

[19] L. Barabási, N. Gulbahce, and J. Loscalzo, “Network medicine: a network-based approach to human disease,” Nature reviews genetics, vol. 12, no. 1, pp. 56–68, 2011.

[20] L. Lin, M. Xiong, G. Zhang, W. Kang, S. Sun, S. Wu, and I. A. D. Neuroimaging, “A convolutional neural network and graph convolutional network based framework for ad classification,” Sensors, vol. 23, no. 4, p. 1914, 2023.

[21] Y. Yin, “Construction and analysis of disease networks based on multiomics data,” Advances in Computer and Communication, vol. 5, no. 6, 2024.

[22] X. Liu, X. Chang, S. Leng, H. Tang, K. Aihara, and L. Chen, “Detection for disease tipping points by landscape dynamic network biomarkers,” National science review, vol. 6, no. 4, pp. 775–785, 2019.

[23] X. Liu, Y. Wang, H. Ji, K. Aihara, and L. Chen, “Personalized characterization of diseases using sample-specific networks,” Nucleic acids research, vol. 44, no. 22, pp. e164–e164, 2016.

[24] N. V. Chawla, K. W. Bowyer, L. O. Hall, and W. P. Kegelmeyer, “Smote: synthetic minority over-sampling technique,” Journal of artificial intelligence research, vol. 16, pp. 321–357, 2002.

[25] W.-D. K. Ma, J. Lewis, and W. B. Kleijn, “The hsic bottleneck: Deep learning without back-propagation,” in Proceedings of the AAAI conference on artificial intelligence, vol. 34, no. 04, 2020, pp. 5085– 5092.

[26] C. Liu, X. Sun, J. Wang, H. Tang, T. Li, T. Qin, W. Chen, and T.-Y. Liu, “Learning causal semantic representation for out-of-distribution prediction,” Advances in Neural Information Processing Systems, vol. 34, pp. 6155–6170, 2021.

[27] Y. Wu, X. Wang, A. Zhang, X. He, and T.-S. Chua, “Discovering invariant rationales for graph neural networks,” in International Conference on Learning Representations, 2022.

[28] Y.-M. He, S. Masuda, T.-B. Jiang, J.-P. Xu, B.-C. Sun, and J.-B. Ge, “Catlet score and clinical catlet score as predictors of long-term outcomes in patients with acute myocardial infarction presenting later than 12 hours from symptom onset,” Annals of medicine, vol. 56, no. 1, p. 2349190, 2024.

[29] D. W. Hosmer Jr, S. Lemeshow, and R. X. Sturdivant, Applied logistic regression. John Wiley & Sons, 2013.

[30] T. Cover and P. Hart, “Nearest neighbor pattern classification,” IEEE transactions on information theory, vol. 13, no. 1, pp. 21–27, 1967.

[31] C. Cortes, “Support-vector networks,” Machine Learning, 1995.

[32] F. Rosenblatt, “The perceptron: a probabilistic model for information storage and organization in the brain.” Psychological review, vol. 65, no. 6, p. 386, 1958.

[33] Z. Liu, Y. Wang, S. Vaidya, F. Ruehle, J. Halverson, M. Soljačić, T. Y. Hou, and M. Tegmark, “Kan: Kolmogorov-arnold networks,” arXiv preprint 2404.19756, 2024.

[34] T. N. Kipf and M. Welling, “Semi-supervised classification with graph convolutional networks,” arXiv preprint 1609.02907, 2016.

[35] P. Veličković, G. Cucurull, A. Casanova, A. Romero, P. Lio, and Y. Bengio, “Graph attention networks,” arXiv preprint 1710.10903, 2017.

[36] W. Hamilton, Z. Ying, and J. Leskovec, “Inductive representation learning on large graphs,” Advances in neural information processing systems, vol. 30, 2017.

[37] K. Xu, W. Hu, J. Leskovec, and S. Jegelka, “How powerful are graph neural networks?” arXiv preprint 1810.00826, 2018.

[38] E. Ranjan, S. Sanyal, and P. Talukdar, “Asap: Adaptive structure aware pooling for learning hierarchical graph representations,” in Proceedings of the AAAI conference on artificial intelligence, vol. 34, no. 04, 2020, pp. 5470–5477.

[39] Z. Ying, D. Bourgeois, J. You, M. Zitnik, and J. Leskovec, “Gnnexplainer: Generating explanations for graph neural networks,” Advances in neural information processing systems, vol. 32, 2019.

[40] S. Shalev-Shwartz and S. Ben-David, Understanding Machine Learning: From Theory to Algorithms. New York, NY, USA: Cambridge University Press, 2014. [Online]. Available: https://www.cambridge.org/core/books/understanding-machine-learning/2A9B85E5BA2B5D866BDBB4C5883E5DC0

[41] P. Lin, Y. Zhang, Z. Chen, J. Chen, G. Chen, J. Fang, Y. Zheng, D. Chen, A. Yidilisi, R. Ji et al., “Interaction between coronary lesion complexity and coronary microvascular dysfunction in the prognosis of nstemi patients,” Clinical Research in Cardiology, pp. 1–13, 2025.

[42] G. De Luca, N. Ernst, F. Zijlstra, A.W. Van’t Hof, J. C. Hoorntje, J.-H. E. Dambrink, A. M. Gosslink, M.-J. de Boer, and H. Suryapranata, “Preprocedural timi flow and mortality in patients with acute myocardial infarction treated by primary angioplasty,” Journal of the American College of Cardiology, vol. 43, no. 8, pp. 1363–1367, 2004.

[43] J. Stamler, R. Stamler, and J. D. Neaton, “Blood pressure, systolic and diastolic, and cardiovascular risks: Us population data,” Archives of internal medicine, vol. 153, no. 5, pp. 598–615, 1993.

[44] P. C. Van den Hoogen, E. J. Feskens, N. J. Nagelkerke, A. Menotti, A. Nissinen, and D. Kromhout, “The relation between blood pressure and mortality due to coronary heart disease among men in different parts of the world,” New England Journal of Medicine, vol. 342, no. 1, pp. 1–8, 2000.

[45] H. Moriyama, T. Kohno, S. Kohsaka, Y. Shiraishi, R. Fukuoka, Y. Nagatomo, A. Goda, A. Mizuno, K. Fukuda, T. Yoshikawa et al., “Length of hospital stay and its impact on subsequent early readmission in patients with acute heart failure: a report from the wet-hf registry,” Heart and vessels, vol. 34, pp. 1777–1788, 2019.

[46] S. Taravatmanesh, D. Khalili, S. Khodakarim, S. Asgari, F. Hadaegh, F. Azizi, and S. Sabour, “Determining the factors associated with cardiovascular disease recurrence: Tehran lipid and glucose study,” The Journal of Tehran University Heart Center, vol. 12, no. 3, p. 107, 2017.

[47] G. Landesberg, V. Shatz, I. Akopnik, Y. G. Wolf, M. Mayer, Y. Berlatzky, C. Weissman, and M. Mosseri, “Association of cardiac troponin, ckmb, and postoperative myocardial ischemia with long-term survival after major vascular surgery,” Journal of the American College of Cardiology, vol. 42, no. 9, pp. 1547–1554, 2003.

[48] S. H. Lee, M. H. Jeong, J. H. Ahn, D. Y. Hyun, K. H. Cho, M. C. Kim, D. S. Sim, Y. J. Hong, J. H. Kim, Y. Ahn et al., “Predictors of recurrent acute myocardial infarction despite successful percutaneous coronary intervention,” The Korean Journal of Internal Medicine, vol. 37, no. 4, p. 777, 2022.

[49] A. O’Neil, A. J. Scovelle, A. J. Milner, and A. Kavanagh, “Gender/sex as a social determinant of cardiovascular risk,” Circulation, vol. 137, no. 8, pp. 854–864, 2018.

[50] V. Mannacio, L. Di Tommaso, V. De Amicis, V. Lucchetti, P. Pepino, F. Musumeci, and C. Vosa, “Previous percutaneous coronary interventions increase mortality and morbidity after coronary surgery,” The Annals of thoracic surgery, vol. 93, no. 6, pp. 1956–1962, 2012.

[51] K. Wita, A. Filipecki, J. Szczogiel, A. Drzewiecka-Gerber, A. Rybicka, J. Krauze, W. Wró bel K. Szydło, D. Urbańczyk, M. Turski et al., “Prediction of adverse cardiac events in patients with acute anterior wall myocardial infarction treated with pci,” Polskie Archiwum Medycyny Wewnetrznej, vol. 116, no. 1, pp. 648–657, 2006.

[52] M. A. Pfeffer, J. J. McMurray, E. J. Velazquez, J.-L. Rouleau, L. Køber, A. P. Maggioni, S. D. Solomon, K. Swedberg, F. Van de Werf, H. White et al., “Valsartan, captopril, or both in myocardial infarction complicated by heart failure, left ventricular dysfunction, or both,” New England Journal of Medicine, vol. 349, no. 20, pp. 1893–1906, 2003.

[53] R. H. Nelson, “Hyperlipidemia as a risk factor for cardiovascular disease,” Primary care, vol. 40, no. 1, p. 195, 2012.

[54] D. Control, C. Trial, E. of Diabetes Interventions, C. D. R. Group et al., “Risk factors for cardiovascular disease in type 1 diabetes,” Diabetes, vol. 65, no. 5, p. 1370, 2016.

[55] P. Amarenco, P. C. Lavallée, J. Labreuche, G. W. Albers, N. M. Bornstein, P. Canhão, L. R. Caplan, G. A. Donnan, J. M. Ferro, M. G. Hennerici et al., “One-year risk of stroke after transient ischemic attack or minor stroke,” New England Journal of Medicine, vol. 374, no. 16, pp. 1533–1542, 2016.

[56] G. J. Hankey, “Long-term outcome after ischaemic stroke/transient ischaemic attack,” Cerebrovascular diseases, vol. 16, no. Suppl. 1, pp. 14–19, 2003.

[57] D. Aune, S. Schlesinger, T. Norat, and E. Riboli, “Tobacco smoking and the risk of heart failure: A systematic review and meta-analysis of prospective studies,” European journal of preventive cardiology, vol. 26, no. 3, pp. 279–288, 2019.

[58] R. Attar, A. Wester, S. Koul, S. Eggert, and P. Andell, “Peripheral artery disease and outcomes in patients with acute myocardial infarction,” Open Heart, vol. 6, no. 1, 2019.

[59] K. Yahagi, H. R. Davis, E. Arbustini, and R. Virmani, “Sex differences in coronary artery disease: pathological observations,” Atherosclerosis, vol. 239, no. 1, pp. 260–267, 2015.

[60] S. Gao, W. Ma, S. Huang, X. Lin, and M. Yu, “Sex-specific clinical characteristics and long-term outcomes in patients with myocardial infarction with non-obstructive coronary arteries,” Frontiers in Cardiovascular Medicine, vol. 8, p. 670401, 2021.

